# Ventricular volume asymmetry as a novel imaging biomarker for disease discrimination and outcome prediction

**DOI:** 10.1101/2023.11.03.23298024

**Authors:** Celeste McCracken, Liliana Szabo, Zaid A. Abdulelah, Hajnalka Vago, Thomas E. Nichols, Steffen E. Petersen, Stefan Neubauer, Zahra Raisi-Estabragh

## Abstract

**Background:** The utility of ventricular asymmetry as an imaging biomarker for cardiovascular risk has not been assessed in population cohorts.

**Objectives:** This study presents a comprehensive assessment of the population distribution of ventricular asymmetry and its relationships across a range of prevalent and incident cardiorespiratory diseases.

**Methods:** Cardiovascular magnetic resonance (CMR) imaging metrics derived from automated image analysis were examined, along with clinical outcomes ascertained through linked health records. Ventricular asymmetry was expressed as the ratio of left and right ventricular (LV, RV) end-diastolic volumes. The normal range for ventricular symmetry was defined in a healthy subset without cardiorespiratory disease. Participants with values outside the 5^th^-95^th^ percentiles of the healthy distribution were classed as either LV dominant (LV/RV > 112%) or RV dominant (LV/RV < 80%) asymmetry. Associations of LV and RV dominant asymmetry with vascular risk factors, CMR features, and prevalent and incident cardiovascular diseases were examined using regression models, adjusting for vascular risk factors, prevalent diseases, and conventional CMR measures.

**Results:** The analysis includes 44,796 participants (average age 64.1±7.7 years; 51.9% women). Ventricular asymmetry, in either direction, was associated with older age and adverse cardiovascular remodeling. LV-dominance was linked to an array of pre-existing vascular risk factors and cardiovascular diseases, and a two-fold increased risk of incident heart failure, non-ischemic cardiomyopathies, and left-sided valvular disorders. RV dominance was associated with an elevated risk of all-cause mortality.

**Conclusions:** Ventricular asymmetry has clinical utility for cardiovascular risk assessment, providing information that is incremental to traditional risk factors and conventional CMR metrics.

**Condensed abstract:** Healthy hearts have a predictable symmetry. Asymmetry produced when one, e.g. the left ventricular (LV) volume outweighs the right, or vice versa, could be an important indicator of underlying disorders, and powerful risk indicator for future disease. In this study of 44,796 UK Biobank participants, we show that LV dominance associates significantly with clinical risk factors, existing heart disease, and a two-fold increased risk for future heart failure, non-ischemic cardiomyopathies, and left-sided valvular disorders. RV dominance was associated with an increased risk of all-cause mortality. Ventricular asymmetry is easily calculated from conventional imaging metrics and could be a highly useful addition to the clinician’s toolkit.

Central illustration:
Ventricular volume asymmetry associates with adverse outcomes

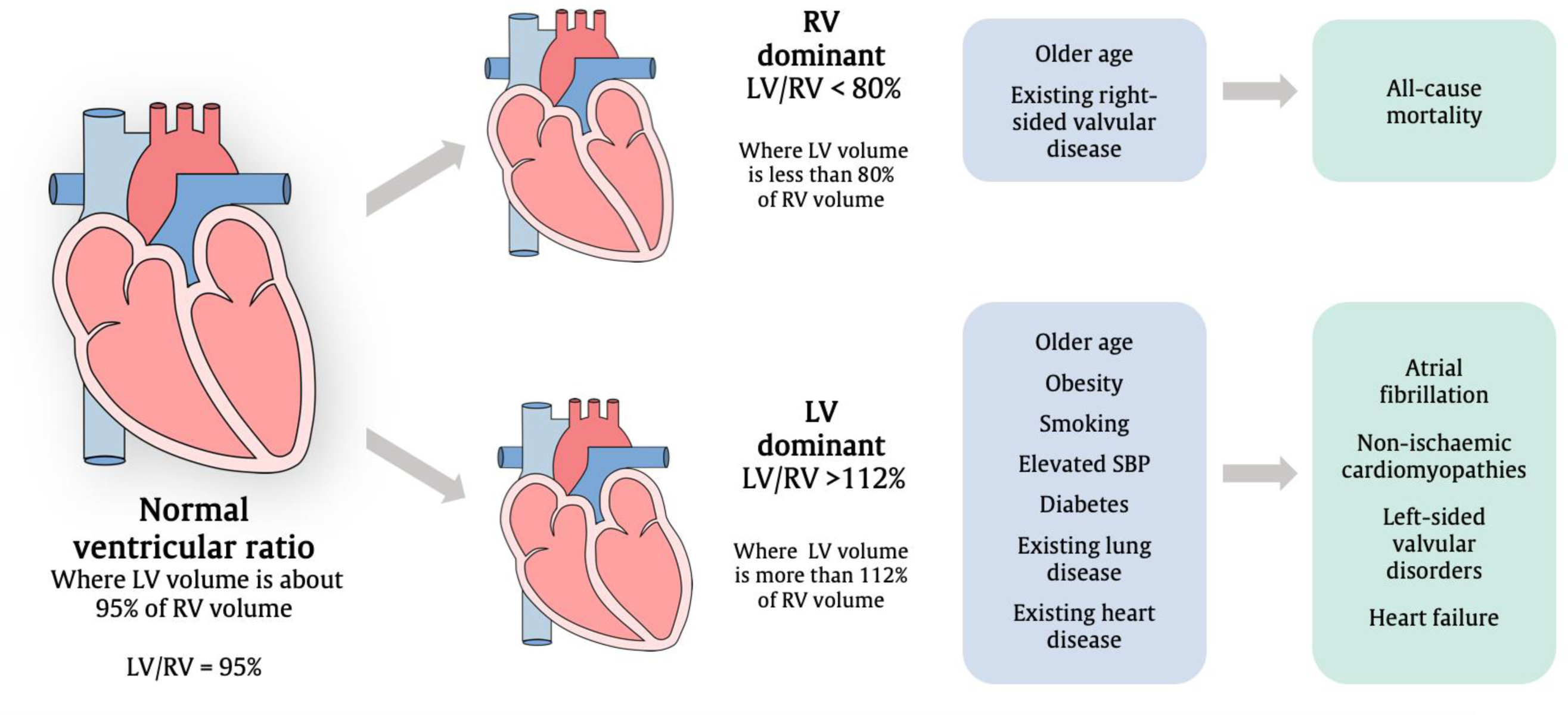

## Introduction

Cardiovascular imaging captures early ventricular remodeling patterns indicative of pre-clinical disease and future cardiovascular risk(1). Left and right ventricular volume measures are routinely used in clinical practice to distinguish health from disease, with dilated ventricles linked to various cardiomyopathies and volume overload states. More recent research indicates that the relative size of the ventricles, in addition to their absolute size, may indicate underlying disease processes(2). In other words, an asymmetric increase in the size of one ventricle over the other may, in itself, be an indicator of cardiovascular risk.

Most of the existing research into the importance of ventricular asymmetry is confined to highly select disease states, where right heart dilatation is prominent. For instance, right-to-left ventricular (RV/LV) volume and diameter ratio have demonstrable value for risk stratification in pulmonary embolism(3), pulmonary artery hypertension(4), and congenital heart disease(5). Furthermore, RV/LV ratio has been used to assess the progression of right ventricular arrhythmogenic cardiomyopathy(2) and to quantify the hemodynamic consequence of pulmonary valve regurgitation in patients with repaired Tetralogy of Fallot(5). The clinical utility of the ventricular ratio metric has not been evaluated outside of these very specific clinical contexts and its utility in wider clinical and population cohorts is unknown. Given that this ratio may be readily calculated from existing routinely available metrics, its potential value as an imaging biomarker merits investigation.

Cardiovascular magnetic resonance (CMR) is the reference modality for volumetric ventricular assessment, providing superior endocardial border definition and yielding highly reproducible biomarkers(6,7). The UK Biobank Imaging Study includes standardized CMR scans for a very large cohort of population-dwelling participants, linked to longitudinally tracked health outcomes, providing the ideal platform to evaluate the value of novel imaging biomarkers for disease discrimination and outcome prediction.

In this study, we examine the associations of left-to-right (LV/RV) and right-to-left (RV/LV) ventricular volume ratios with a wide range of prevalent and incident cardiovascular risk factors and diseases in a large sample of UK Biobank participants. Importantly, we evaluate the incremental value of the ventricular ratio metric over clinical risk factors and existing conventional measures of ventricular and atrial function.

## Methods

### Study population

The UK Biobank is a prospective cohort study of half a million participants from a range of urban and rural settings across the UK. Individuals aged 40 to 69 years old living within 25 miles of 22 UK Biobank assessment centers were recruited from National Health Service registers between 2006 and 2010, where extensive baseline phenotyping was conducted(8). The UK Biobank Imaging Study was launched in 2015 and aims to scan a 20% (n=100,000) subset of the original participants using a standardized acquisition protocol for multiorgan imaging, which includes CMR(9). Extensive health record linkage has been established for the entire UK Biobank cohort permitting longitudinal tracking of incident health events. For this study, we included 44,796 participants with imaging metrics available with scanning taking place between April 2014 and March 2020 (**Supplementary Figure 1**).

### Ethics

This study complies with the Declaration of Helsinki; the work was covered by the ethical approval for UK Biobank studies from the NHS National Research Ethics Service on 17th June 2011(Ref 11/NW/0382) and extended on 18th June 2021(Ref 21/NW/0157) with written informed consent obtained from all participants.

### CMR protocol and image analysis

CMR imaging was performed on 1.5 Tesla scanners (MAGNETOM Aera, Syngo Platform VD13A, Siemens Healthcare, Erlangen, Germany) in dedicated imaging centers using standardized staff training and equipment, as described in a dedicated publication(10). Image analysis was conducted with automated pipelines with in-built quality control validated in the UK Biobank, as described in previous work(9,11–13). For this study, the following CMR metrics were included: left ventricular end-diastolic volume index (LVEDVi), left ventricular mass index (LVMi), maximal wall thickness (Max WT), myocardial native T1, left ventricular ejection fraction (LVEF), left ventricular global function index (LVGFI), global longitudinal strain (GLS), global circumferential strain (GCS), right ventricular end diastolic volume index (RVEDVi), right ventricular ejection fraction (RVEF), left atrial ejection fraction (LAEF), right atrial ejection fraction (RAEF) and aortic distensibility (AoD).

### Definition of LV and RV dominant ventricular asymmetry

We calculated left-to-right (LV/RV) and right-to-left (RV/LV) ventricular ratios using LVEDVi and RVEDVi. These two ratios are not interchangeable and could provide distinct insights into cardiac function. **Figure 1** illustrates the mathematical relationship between left-to-right (LV/RV) and right-to-left (RV/LV) ventricular ratios.

**Figure 1:**
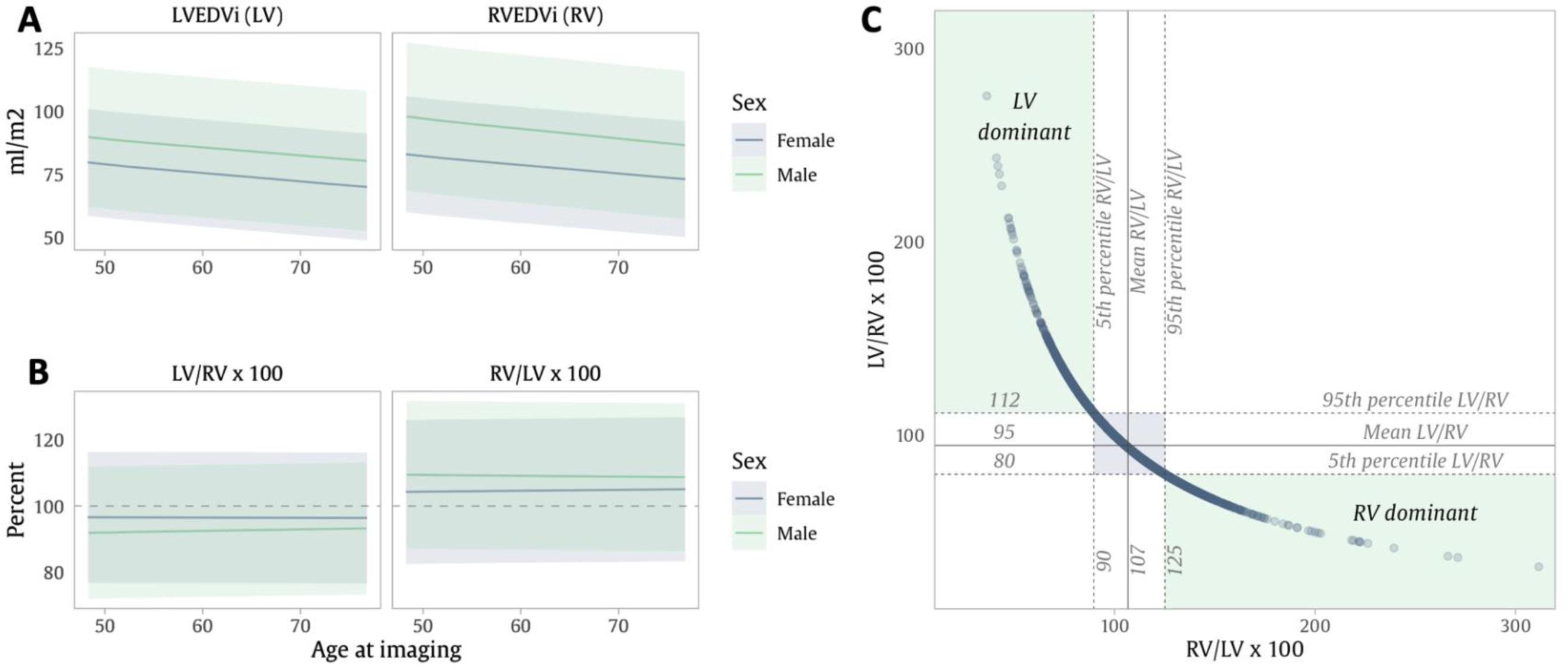
Ventricular volume symmetry in a healthy subset of participants free from clinical cardiorespiratory disease (n= 32,700) **A**: Average values for left and right end-diastolic volume indexed to body surface area by age in the healthy subset, with men in green and women in blue. Shaded areas indicate the 95% prediction interval for each sex. **B**: Average ventricular asymmetry ratios by age in the healthy subset, with men in green and women in blue. Shaded areas indicate the 95% prediction interval for each sex. **C**: The distribution of ventricular asymmetry in the healthy subset, with the areas of asymmetry shown in green. The shaded area in blue indicates the middle symmetry range comprising 90% of the healthy subset. Abbreviations: LV: left ventricle; LVEDVi: LV end-diastolic volume indexed to body surface area; RV: right ventricle; RVEDV: RV end-diastolic volume indexed to body surface area.

To understand the normal variations of ventricular asymmetry, we created a subset of 37,200 participants free from clinically diagnosed cardiorespiratory disease (as per linked health records or UK Biobank assessments) at the time of imaging (**Supplementary Figure 1**). This subset was used to define the average normal ventricular symmetry for men and women, along with their 5^th^ and 95^th^ percentiles (**Table 1** and **Supplementary Table 1**). **Figure 1C** demonstrates the 5^th^ and 95^th^ percentiles, indicating the well-defined range within which 90% of healthy individuals fall. Outside this range, we observe two types of ventricular asymmetry: cases where the LV is disproportionately larger than the RV (LV/RV > 112%), labeled as “LV dominant”, and cases where the RV is significantly larger relative to the LV (LV/RV < 80%), labelled as “RV dominant”. To simplify the associations with ventricular asymmetry, we created binary variables for abnormal ventricular ratios, flagging participants with LV or RV dominant asymmetry, which allowed for a more nuanced understanding of the exposure to ventricular volume imbalance. Following exploratory analysis, we added a third category for mildly asymmetric cases, where LV/RV is not abnormal, but is more than 1 standard deviation (SD) above the healthy subset mean. We call this category “mildly LV dominant”.

**Table 1:**
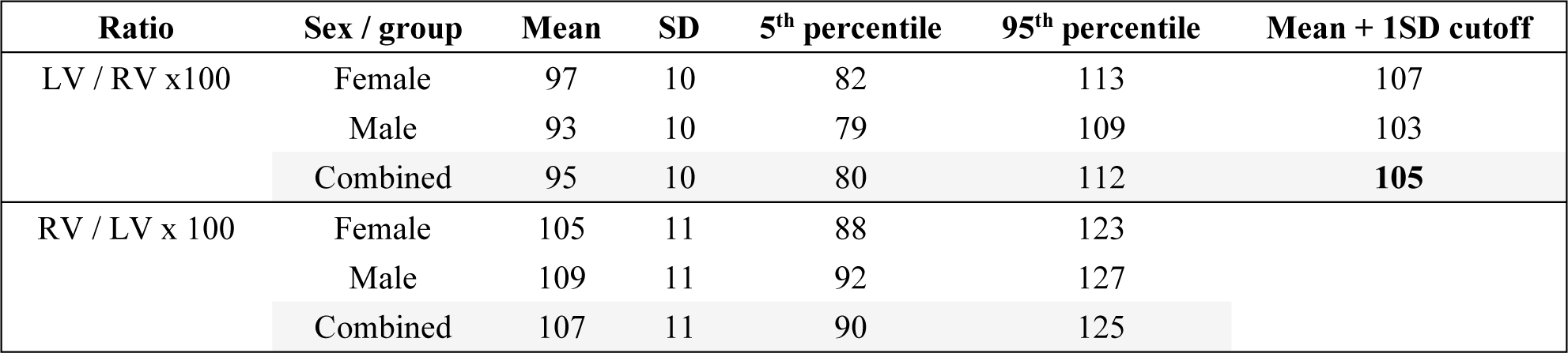
Normal range of ventricular symmetry in the healthy subset (n= 32,700)

### Definition of covariates

Sex, ethnicity, family history of heart disease, and Townsend deprivation index were recorded at baseline recruitment. Age, body mass index (BMI), waist-to-hip ratio, and systolic blood pressure (SBP) were assessed at the imaging visit. Smoking, alcohol intake frequency, and physical activity were self-reported at the imaging visit. Hypertension, diabetes, and high cholesterol status at imaging were ascertained by both self-report and linked hospital records (**Supplementary Table 2**).

### Ascertainment of outcomes

Prevalent conditions existing at the time of imaging were ascertained by both self-report and linked hospital records. Incident diagnoses – occurring for the first time after imaging – were indicated by linked hospital records only (**Supplementary Table 2**). We recorded stroke (any type), myocardial infarction, atrial fibrillation, non-ischemic cardiomyopathies, heart failure, right-sided valvular disorders (tricuspid or pulmonary), left-sided valvular disorders (mitral or aortic), any cardiovascular disease (CVD, meaning any ICD10 code between I00-I80), CVD mortality (death where the primary cause is between I00-I80) and all-cause mortality. We also included several respiratory diseases to report potential asymmetry at imaging, namely asthma, chronic obstructive pulmonary disease (COPD), obstructive sleep apnea, interstitial lung disease, and bronchiectasis. The study censor date was October 2022, providing an average prospective follow-up time of 4.75 ± 1.52 years. Participants with records of the disease of interest prior to imaging were excluded from modeling for incidence of that outcome.

### Statistical analysis

Analysis was performed using R version 4.2.3 and RStudio Version 2023.03.0+386. We explored the relationships of each ventricular asymmetry type (RV dominant, LV dominant, mildly LV dominant) with a selection of clinical and imaging indicators of cardiorespiratory health. We used unadjusted logistic regression to describe the odds of each asymmetry type associated with clinical risk factors such as elevated SBP (> 140mmHg), physical activity, and smoking.

To understand associations between asymmetry and other CMR metrics, we employed a series of linear regression models with asymmetry entered as a binary exposure variable (no asymmetry set as the reference level) and other CMR metrics inserted in turn as the continuous outcome variable. The models were adjusted for age, sex, SBP, BMI, deprivation, hypertension, high cholesterol, and diabetes. Standardized betas were reported for each association showing the SD difference in each CMR metric associated with asymmetry status.

Associations with existing disease at imaging were examined using multivariable logistic regression models with asymmetry category (separately for RV dominant, LV dominant, and mildly LV dominant) set as the outcome, and existing disease as the predictor, adjusted by age, sex, SBP, BMI, and deprivation. Therefore, the odds ratio shows the adjusted odds of having each ventricular asymmetry type given existing cardiorespiratory disease.

Finally, we examined the association of each asymmetry type with incident cardiovascular events using Cox proportional hazard models. Our analysis encompassed three layers of adjustment: 1) Unadjusted associations between LV and RV dominant ventricular asymmetry and incident cardiovascular events. 2) Demographic and clinical adjustment: models adjusted for age, sex, SBP, BMI, deprivation, hypertension, high cholesterol, and diabetes. 3) Fully adjusted models, including model 2 covariates plus additional adjustment for selected CMR metrics that have been linked to adverse cardiovascular outcomes in the literature - LVEF, RVEF and LVMi. Results are presented as hazard ratios for incident disease associated with the presence of asymmetry alongside 95% confidence intervals and p-values. Multiple testing correction is applied across all significance thresholds using a 5% false discovery rate.

## Results

### Baseline characteristics

The analysis includes 44,796 participants, including 21,545 (48.1%) men and 23,251 (51.9%) women with average age of 64.1 (±7.7) years (**Table 2**). Within the sample, 33.8% had hypertension, 29.3% had high cholesterol, and 5.9% had diabetes. Overall, 2,263 (5%) had RV dominant asymmetry, 2,658 (6%) had LV dominant asymmetry, and 6,656 (15%) participants had mildly LV dominant asymmetry. Over the follow-up period, we observed 2,239 new cases of CVD (**Supplementary Table 3**), and 785 cases of all-cause mortality. Atrial fibrillation was the most commonly observed incident diagnosis with 958 cases, and right-sided valvular disorder was the rarest (81 cases). Means and standard deviations for all CMR metrics measured in all symmetry groups are provided in **Supplementary Table 4**.

**Table 2:** Sample characteristics.

| Characteristic | Whole sample<br>(n= 44,796) | Within normal symmetry<br>(n= 39,875) | RV dominant<br>(n= 2,263) | LV dominant<br>(n= 2,658) |
| --- | --- | --- | --- | --- |
| Age (years) | 64.1 ($\pm 7.7$) | 63.9 ($\pm 7.7$) | 65.8 ($\pm 7.7$) | 66.5 ($\pm 7.4$) |
| Women | 23,251 (51.9%) | 20,781 (52.1%) | 1,185 (52.4%) | 1,285 (48.3%) |
| Men | 21,545 (48.1%) | 19,094 (47.9%) | 1,078 (47.6%) | 1,373 (51.7%) |
| Non-white ethnicity | 1,336 (3.0%) | 1,201 (3.0%) | 72 (3.2%) | 63 (2.4%) |
| Townsend deprivation index | -2.6 [-3.9, -0.4] | -2.6 [-3.9, -0.4] | -2.5 [-3.8, -0.5] | -2.6 [-3.9, -0.4] |
| Above median UK deprivation (more deprived) | 10,882 (24.3%) | 9,677 (24.3%) | 543 (24.0%) | 662 (24.9%) |
| Body mass index (kg/m<sup>2</sup>) | 25.9 [23.5, 28.8] | 25.9 [23.5, 28.8] | 25.7 [23.3, 28.3] | 25.9 [23.4, 28.9] |
| Obesity (BMI > 30 kg/m<sup>2</sup>) | 8,028 (17.9%) | 7,149 (17.9%) | 356 (15.7%) | 523 (19.7%) |
| Waist-hip ratio > 1 | 3,039 (6.8%) | 2,640 (6.6%) | 168 (7.4%) | 231 (8.7%) |
| Systolic blood pressure (mmHg) | 139.0 ($\pm 18.7$) | 138.7 ($\pm 18.5$) | 136.7 ($\pm 17.7$) | 145.5 ($\pm 20.8$) |
| Alcohol intake frequency<sup>1</sup> |  |  |  |  |
| Never | 2,963 (6.6%) | 2,583 (6.5%) | 161 (7.1%) | 219 (8.2%) |
| Less than once per week | 9,682 (21.6%) | 8,631 (21.6%) | 506 (22.4%) | 545 (20.5%) |
| Once per week or more | 31,821 (71.0%) | 28,373 (71.2%) | 1,574 (69.6%) | 1,874 (70.5%) |
| Smoking<sup>1</sup> |  |  |  |  |
| Never smoker | 27,780 (62.0%) | 24,854 (62.3%) | 1,432 (63.3%) | 1,494 (56.2%) |
| Previous smoker | 15,181 (33.9%) | 13,400 (33.6%) | 748 (33.1%) | 1,033 (38.9%) |
| Current smoker | 1,516 (3.4%) | 1,344 (3.4%) | 61 (2.7%) | 111 (4.2%) |
| Physically active<sup>2</sup> | 37,504 (83.7%) | 33,489 (84.0%) | 1,857 (82.1%) | 2,158 (81.2%) |
| Ventricular volume ratios |  |  |  |  |
| LV/RV * 100 | 94.5 [88.3, 101.1] | 94.4 [88.9, 100.1] | 76.9 [74.3, 78.9] | 116.5 [113.5, 122.3] |
| RV/LV * 100 | 106.3 ($\pm 11.7$) | 106.3 ($\pm 8.5$) | 132.5 ($\pm 11.6$) | 83.9 ($\pm 6.9$) |
| Hypertension | 15,130 (33.8%) | 13,150 (33.0%) | 741 (32.7%) | 1,239 (46.6%) |
| High cholesterol | 13,110 (29.3%) | 11,381 (28.5%) | 693 (30.6%) | 1,036 (39.0%) |
| Diabetes | 2,624 (5.9%) | 2,274 (5.7%) | 132 (5.8%) | 218 (8.2%) |
| Family history of heart disease | 24,563 (54.8%) | 21,819 (54.7%) | 1,502 (56.5%) | 1,242 (54.9%) |

### Ventricular symmetry in the healthy subset

Within the 32,700 participants in the healthy subset without cardiorespiratory disease at the time of imaging, the indexed left and right ventricular volumes were larger in men, and declined with increasing age (**Figure 1A**). In contrast, average ventricular ratio (LV/RV and RV/LV) were very similar in men and women and did not exhibit significant age dependency (**Figure 1B** and **Supplementary Table 1**).

### Associations between ventricular asymmetry and clinical factors

Participants aged 70 or over were significantly more likely to have ventricular asymmetry of both types (**Figure 2** and **Supplementary Table 5**). LV dominant asymmetry was seen more frequently in participants with BMI > 30 kg/m^2^, waist-to-hip ratio greater than 1.0, current and previous smoking, and SBP above 140 mmHg. Participants who were physically active (greater than 600 summed MET minutes per week) were less likely to have ventricular asymmetry in either direction.

**Figure 2:**
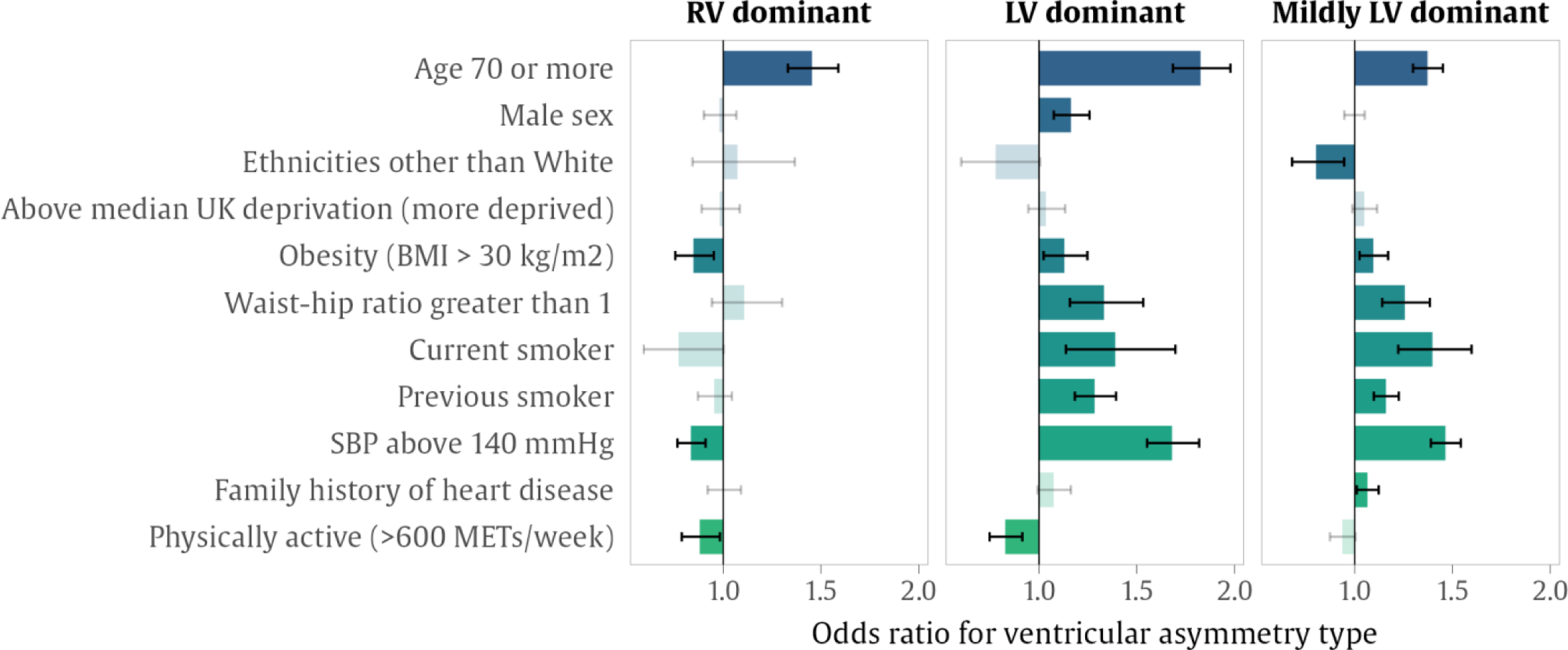
Associations between ventricular asymmetry and risk factors. Bars show odds ratio for ventricular asymmetry associated with the presence of each risk factor, calculated with logistic regression. Intervals show the 95% confidence interval for the odds ratio, where two-tailed significance was adjusted with a false discovery rate of 5%. Where associations were not significant, these are shown in greyed-out color. Abbreviations: BMI: body mass index; LV: left ventricle; MET: metabolic equivalents; RV: right ventricle; SBP: systolic blood pressure.

### Associations between ventricular asymmetry and CMR metrics

Compared to those with ventricular symmetry in the normal range, LV dominant asymmetry was associated a consistent pattern of adverse cardiovascular phenotypic alterations (**Figure 3** and **Supplementary Table 6**). This was characterized by dominant associations with unhealthy myocardial remodeling (higher LVMi, greater maximal WT, higher myocardial native T1) and worse LV function (lower LVEF, lower LVGFI, deteriorating GLS and GCS). LV dominant asymmetry was also linked to reduced RV and LA function (lower RVEF and LAEF) and declining aortic compliance (lower AoD). Notably, these adverse associations also appeared significant (but of smaller magnitude) in the group with mildly LV dominant asymmetry (LV/RV greater than one standard deviation above the healthy mean).

**Figure 3:**
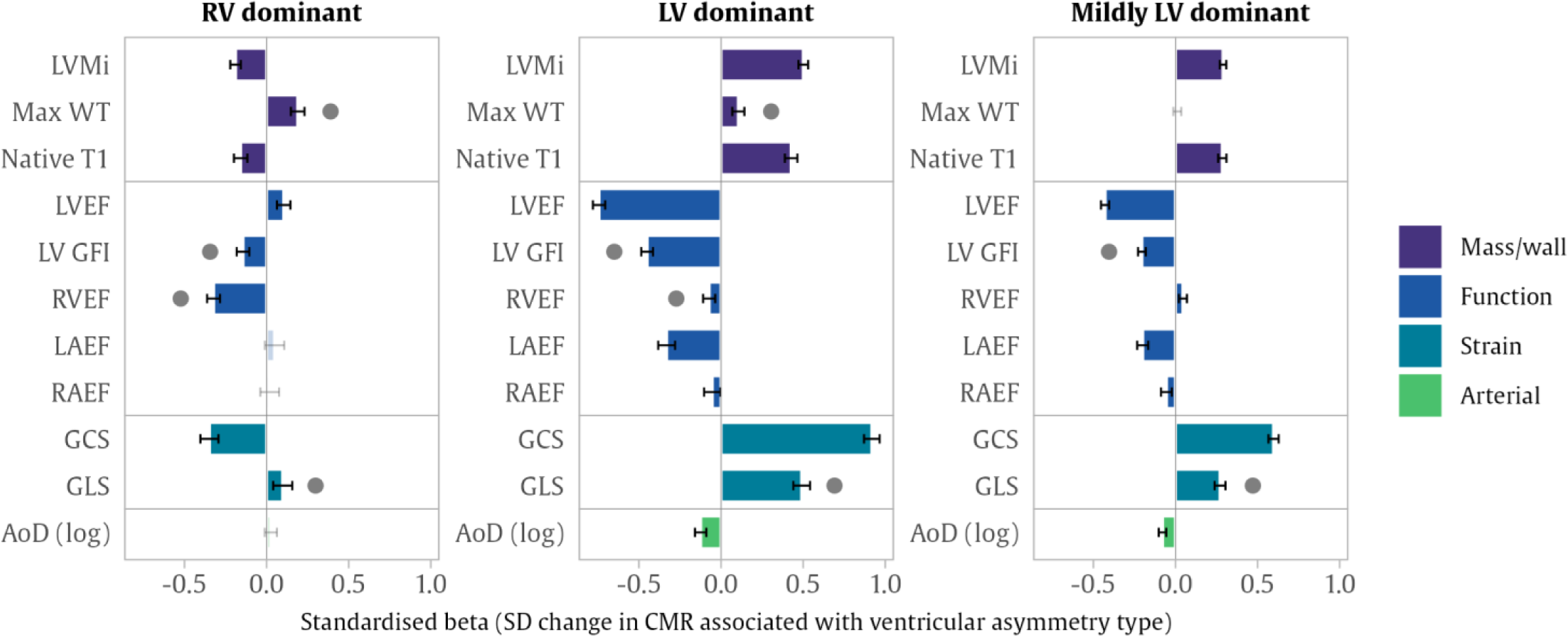
Associations between ventricular asymmetry and other CMR metrics. Bars show standardised beta coefficients for differences in CMR measures associated with each type of ventricular asymmetry compared to hearts within the normal symmetry range. Each bar is from a separate linear regression model, relating symmetry category to CMR metric, adjusted by age, sex, body mass index, systolic blood pressure, smoking, Townsend deprivation index, clinically diagnosed hypertension, diabetes and high cholesterol. Error bars indicate the 95% confidence interval, with two-tailed significance adjusted with a false discovery rate of 5%. Grey dots indicate significant CMR alterations that are found in asymmetry in either direction. Abbreviations: LVMi: Left ventricular mass indexed to body surface area; WT: wall thickness; LVEF: left ventricular ejection fraction; LV GFI: left ventricular global function index; RVEF: right ventricular ejection fraction; LAEF: left atrial ejection fraction; RAEF: right atrial ejection fraction; GCS: global circumferential strain; GLS: global longitudinal strain; AoD: aortic distensibility.

RV dominant asymmetry was associated with significantly lower RVEF, poorer LV function by GLS and LVGFI, and greater maximal WT. The magnitude of the RVEF effect was much larger in RV dominant than LV dominant asymmetry. RV dominance was linked to paradoxically higher LVEF and lower myocardial native T1. Mixed effects were observed in the relationships of RV dominant asymmetry with LV strain parameters, with higher (worse) GLS values but a decrease (apparently healthier) in GCS.

### Associations between ventricular asymmetry and prevalent disease

LV dominant asymmetry was strongly associated with pre-existing cardiac and respiratory disease, notably myocardial infarction, non-ischemic cardiomyopathies, left-sided valvular disorders (mitral or aortic), heart failure, interstitial lung diseases, and COPD (**Figure 4** and **Supplementary Table 7**). These associations were also significant in the group with mild LV asymmetry. On the other hand, compared with symmetric hearts, participants with RV dominant asymmetry were three times more likely to also have existing right-sided valvular disorders (tricuspid and pulmonary).

**Figure 4:**
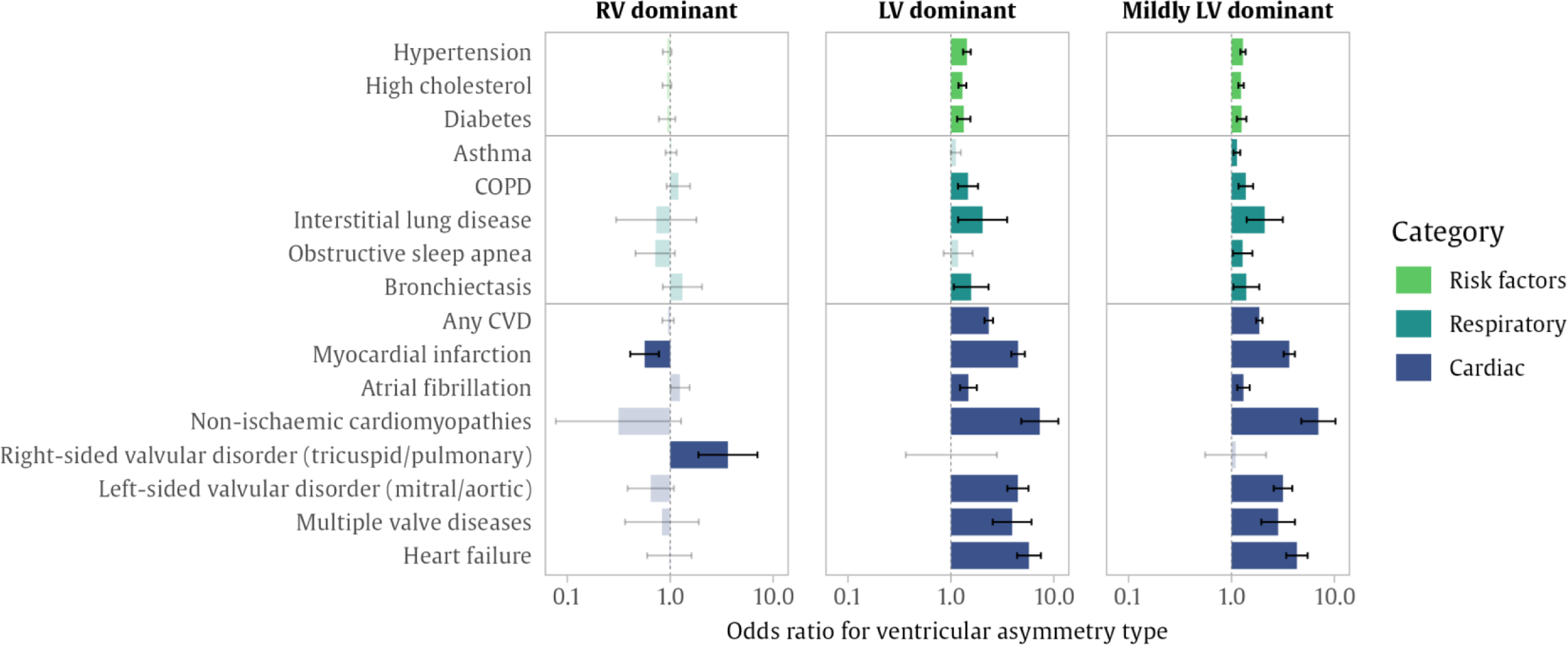
Associations between existing disease and ventricular asymmetry. Bars show odds ratio for ventricular asymmetry associated with the presence of each existing diagnosis, calculated with logistic regression, and adjusted by age, sex, body mass index, systolic blood pressure and Townsend deprivation index. Intervals show the 95% confidence interval for the odds ratio, where two-tailed significance was adjusted with a false discovery rate of 5%. Where associations were not significant, these are shown in greyed-out color. Abbreviations: CVD: cardiovascular disease; COPD: chronic obstructive pulmonary disease.

### Associations between ventricular asymmetry and incident adverse cardiovascular events

LV dominant asymmetry as a standalone measurement demonstrated a strong association with increased risk across all outcomes considered except right-sided valvular disorders (**Figure 5** and **Supplementary Table 8**). In fully-adjusted models, which accounted for a wide range of demographic and clinical covariates and conventional CMR measures, LV dominance was associated with an increased risk for incident atrial fibrillation and a more than two-fold increased risk for heart failure, non-ischemic cardiomyopathies, and left-sided valvular disorders. Importantly, these associations persisted in the group of participants with mild LV dominant asymmetry, with an additional association with myocardial infarction. While RV dominant asymmetry was not associated with any of the incident diseases considered, it was associated with significantly greater risk of all-cause mortality.

**Figure 5:**
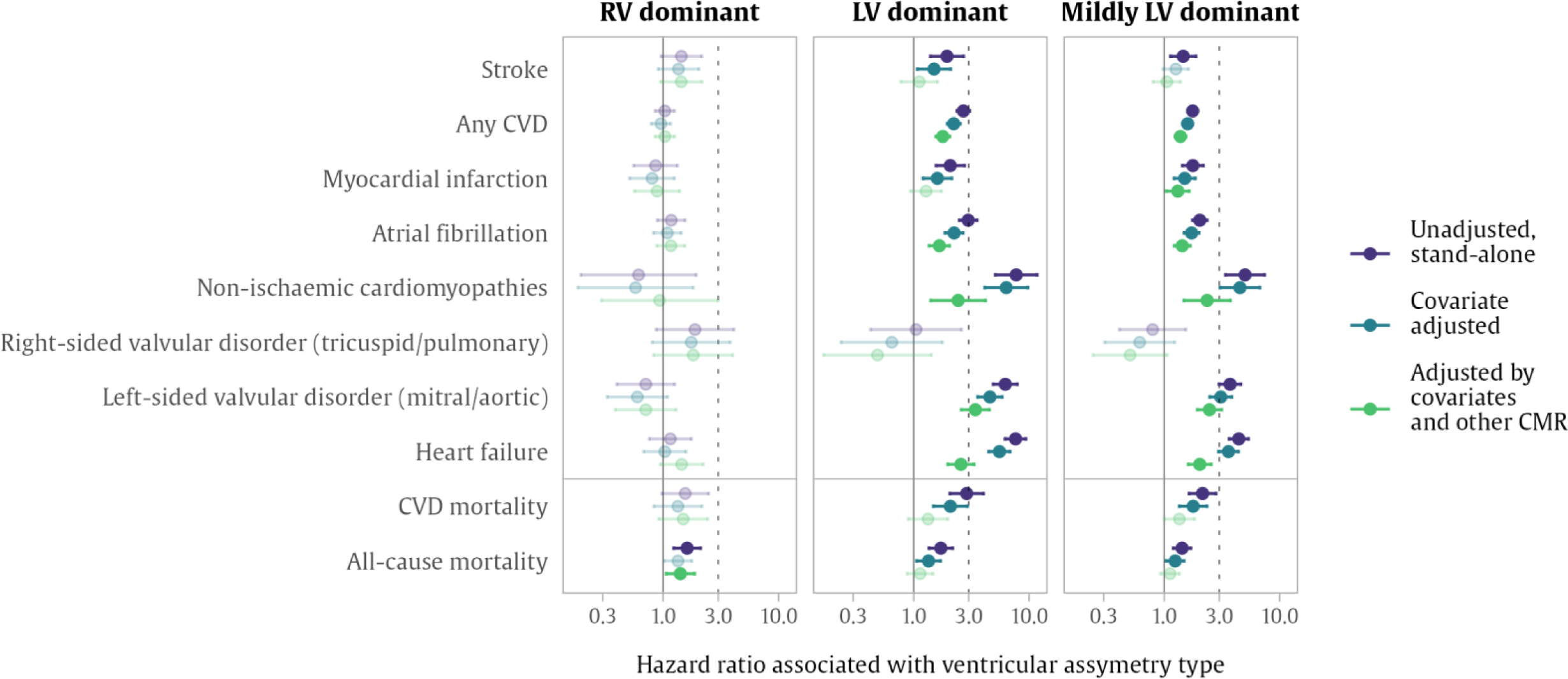
Associations between ventricular asymmetry and incident cardiovascular disease. Dots are hazard ratios for each disease associated with each volume asymmetry type, with 95% confidence intervals Cox proportional hazard regression models. The vertical dotted line indicates a 3-fold hazard. Model 1 (in purple) are unadjusted associations. Model 2 (in blue) are adjusted by age, sex, systolic blood pressure, body mass index, smoking, Townsend deprivation index, hypertension, high cholesterol, and diabetes. Model 3 (in green) are adjusted by Model 2 covariates plus left ventricular mass index, right ventricular ejection fraction and left ventricular ejection fraction. Associations that are not significant are shown in greyed-out color, where the two-tailed significance was adjusted for multiple testing with a false discovery rate of 5%. Abbreviations: CMR: cardiovascular magnetic resonance; CVD: cardiovascular disease; LV: left ventricle; RV: right ventricle.

## Discussion

### Summary of findings

This study of 44,796 UK Biobank participants characterizes the distribution and clinical associations of CMR-derived ventricular asymmetry and examines the incremental clinical utility of this metric for risk prediction over established clinical and imaging indicators.

In healthy participants, average ventricular symmetry was steady across all ages, while absolute ventricular volumes exhibited significant age dependency. In the overall sample, there was greater ventricular asymmetry (in both directions) with older age.

LV dominant asymmetry emerged as a reliable indicator for an extensive range of prevalent vascular risk factors, pre-existing cardiorespiratory diseases, and adverse cardiovascular remodeling. Specifically, LV dominant asymmetry demonstrated significant associations with adverse metabolic profile, prevalent ischemic and non-ischemic heart diseases, pre-existing obstructive and interstitial lung diseases, and a remodeling pattern indicative of pathologic myocardial alterations (increased max WT, increased LVMi, increased T1), cardiac dysfunction (LV, RV, LA) by various volumetric and strain metrics, and poorer aortic compliance (lower AoD). Importantly, LV dominant asymmetry was associated with significantly increased risk of incident CVDs (atrial fibrillation, non-ischemic cardiomyopathies, heart failure, left-sided valve diseases) independent of clinical risk factors and conventional CMR measures with established prognostic importance (LVEF, RVEF, LVMi). These relationships were not confined to those with extreme LV-asymmetry, and a similar pattern of associations was observed in individuals with mildly LV dominant asymmetry (1 SD above the healthy subset average). RV dominant asymmetry showed dominant associations with aging, reduced RV function (lower RVEF), and prevalent right-sided valve disorders. RV dominant asymmetry was associated with increased risk of all-cause mortality independent of established risk factors and CMR metrics.

### Comparison with existing literature

Ventricular volume ratio, as a measure of ventricular asymmetry, represents a simple and readily calculable imaging biomarker for cardiovascular risk assessment, derived from existing ventricular measurements. Prior literature proposes that RV dominant asymmetry, assessed by either RV/LV volume or diameter ratio plays a significant role in the risk stratification of specific conditions such as pulmonary embolism and pulmonary hypertension(14–17). Our study provides new insights by examining associations of both LV and RV dominant asymmetry and by considering relationships in a population sample without selection on any disease. While we corroborate previously reported associations of RV dominant asymmetry with all-cause death, we found that, in our sample, LV dominant asymmetry exerted a more significant impact on incident cardiovascular outcomes. This observation is not surprising and reflects the greater burden of left-sided heart diseases in the general population, compared to previous studies which had focused on less common diseases with prominent RV remodeling. Our findings significantly extend the remit of ventricular asymmetry beyond the limited conditions proposed in previous research and highlight the incremental prognostic value of LV dominant asymmetry in the general population over established clinical and imaging indicators.

Our study paints a vivid picture of the adverse cardiovascular remodeling pattern associated with LV dominant asymmetry, independent of clinical risk factors. Cardiac remodeling, alterations in the structure and function of the heart and vasculature in response to stressors, often precedes the occurrence of clinically detectable disease and is a critical component of the progression of many CVDs. Our analysis demonstrates significant relationships of both abnormal and mild LV dominant asymmetry with a pathologic pattern of cardiovascular remodeling as indicated by greater LV hypertrophy (higher LVMi, greater max WT), greater myocardial fibrosis (higher native), and poorer contractility by volumetric (lower LVEF, lower LVGFI) and strain (worse GLS and GCS) metrics. While the associations with LV remodeling were most notable, LV dominant asymmetry also associated with worse RV and LA (lower RVEF, lower LAEF) function and greater aortic stiffness (lower AoD). Our findings indicate that LV dominant asymmetry presents a reflection of a range of maladaptive, potentially subclinical, cardiac remodeling. The pathway driving the relationship between LV dominant asymmetry and cardiac remodeling is multifaceted. Chronic pressure overload in the systemic circulation could be a potential driver, instigating LV hypertrophy and consequential tissue level modifications (18). Additionally, our study noted an association of LV dominant asymmetry with elevated T1 values, a surrogate marker for diffuse myocardial fibrosis (19). Fibrosis of the myocardium can cause an increase in ventricular stiffness, subsequently leading to systolic and diastolic dysfunction and ultimately increased risk of heart failure and cardiac-related mortality.

Moving beyond cardiac remodeling patterns, we further elucidated associations between LV dominant asymmetry and prevalent cardiometabolic risk factors, including older age and higher BMI. Both aging and obesity are well-documented risk factors for cardiovascular disease, with their impact manifesting through a series of pathophysiological changes including hypertension, diabetes, and dyslipidemia, together or independently(20). Given that these conditions primarily impact the systemic circulation and the LV, resultant cardiovascular remodeling will preferentially impact the LV and potentially lead to the development of an LV-dominant direction of ventricular asymmetry.

Moreover, our study provides compelling evidence that LV dominant asymmetry is significantly associated with both prevalent and future cardiorespiratory disease, even after adjusting for conventional imaging-derived biomarkers such as LVEF or LVMi. This observation adds a prognostic dimension to our understanding of ventricular asymmetry, underscoring its potential value as a risk stratification tool. This also suggests that the LV dominant asymmetry provides additional prognostic information beyond traditional measures of cardiac function. As an example, in heart failure the association with LV dominance appears to be multifactorial with one of the leading factors being the activation of the renin-angiotensin-aldosterone system which provokes significant left ventricular remodeling(21). LV dominant asymmetry was strongly associated with the development of AF with that being elucidated by LV dominance inducing left atrial remodeling driven by progressive accumulation of extracellular matrix in the atrial tissue which consequently contributes towards atrial enlargement(22,23). Notably, these associations were also observed in our study even when among those with mildly LV dominant asymmetry (ventricular asymmetry in the normal range but 1 SD above the healthy subset mean), emphasizing the clinical importance of even subtle shifts in this ratio. However, the detailed molecular and cellular mechanisms that connect ventricular asymmetry to cardiac remodeling and disease development warrant further investigation. Unraveling these underlying mechanisms could lead to the identification of novel therapeutic targets and further enhance our ability to manage and prevent cardiorespiratory diseases.

Our study found a significant association between RV dominant remodeling and an increased risk of all-cause mortality. Importantly, this association was seen in the absence of an associated increase in most prevalent or incident cardiovascular disease. While an association with prevalent right-sided valvular disease was observed, its rarity makes it unlikely to be the primary explanation for the increased mortality seen in our study population. Instead, the observed RV remodeling may be a more global indicator of systemic or non-cardiac conditions that contribute to increased all-cause mortality. Indeed, adverse RV remodeling has been previously associated with systemic conditions, including liver disease (24,25), renal disease (26), and cancer (27), indicating a potential multisystem involvement that could influence overall patient health and mortality risk. Furthermore, biological processes like inflammation, oxidative stress, or fibrosis, all of which have been linked with RV remodeling (28), could also be contributing to this increased mortality. This supports the idea that RV remodeling could be a surrogate marker of these underlying processes, which have systemic implications beyond the cardiovascular system.

### Clinical implications

The observations from our study offer noteworthy clinical implications. The evaluation of ventricular asymmetry through simple volumetric assessment, as performed in this work, presents an opportunity to delve deeper into disease pathogenesis and progression. The observed associations between ventricular asymmetry, specifically LV dominant asymmetry, and adverse cardiovascular outcomes suggest its potential as a sensitive marker for early or subclinical disease stages. This utility is further emphasized by its prognostic value independent of conventional imaging biomarkers such as LVEF. Given that ventricular asymmetry can be evaluated through simple volume ratio assessment as performed in our study, its integration into clinical routine would not require significant additional resource investment. By enabling early detection and intervention in at-risk individuals, this could potentially lead to substantial long-term savings, however formal cost-effectiveness studies are needed to confirm these potential benefits.

### Study limitations

The UK Biobank data regarding incident disease and outcomes were derived from linked hospital records and thus do not include any incident outcomes identified in an outpatient setting. It is noteworthy that despite the large number of enrolled participants, the UK Biobank is widely regarded to lack the appropriate representativeness of the population in a variety of sociodemographic and participant-specific factors as it has been previously reported(29). Despite that, the established heterogeneity of the UK Biobank provides a credible source for scientific identification of associations between metrics and cardiovascular diseases that can be generalized towards other populations(29). It is also important to note that with older automated tools, the measurement of RV volume can be less reproducible than LV volume, due to its challenging presentation and mechanics(30), therefore careful consideration of this issue is key when constructing LV/RV ratios in practice.

### Conclusions

In conclusion, the observations from this large-scale imaging study enrich our understanding of cardiovascular remodeling and disease progression. Our study underscores the significant association between ventricular asymmetry, specifically LV dominant asymmetry, and prevalent as well as incident cardiorespiratory diseases, and its prognostic value. The evaluation of ventricular asymmetry offers an important opportunity to detect early or subclinical stages of disease, which can aid in early intervention and potential improvement of patient outcomes. As this assessment can be performed using simple volumetric measurements, it presents a cost-effective and feasible addition to routine clinical practice. The detailed biological pathways and mechanisms that underpin these associations need further elucidation.

## Data Availability

This research was conducted using the UK Biobank resource under access application 59867. UK Biobank will make the data available to all bona fide researchers for all types of health-related research that is in the public interest, without preferential or exclusive access for any persons. All researchers will be subject to the same application process and approval criteria as specified by UK Biobank. For more details on the access procedure, see the UK Biobank website: https://www.ukbiobank.ac.uk/enable-your-research/register

## Acknowledgements

This study was conducted using the UK Biobank resource under access application 2964. We would like to thank all the participants, and staff involved with planning, collection and analysis, including core lab analysis of the CMR imaging data. This work uses data provided by patients and collected by the NHS as part of their care and support, with copyright © (2023), NHS Digital. Re-used with the permission of the NHS Digital [and/or UK Biobank]. All rights reserved. This research used data assets made available by National Safe Haven as part of the Data and Connectivity National Core Study, led by Health Data Research UK in partnership with the Office for National Statistics and funded by UK Research and Innovation (grant ref MC_PC_20058).

## Supplementary Materials

**Supplementary Figure 1:**
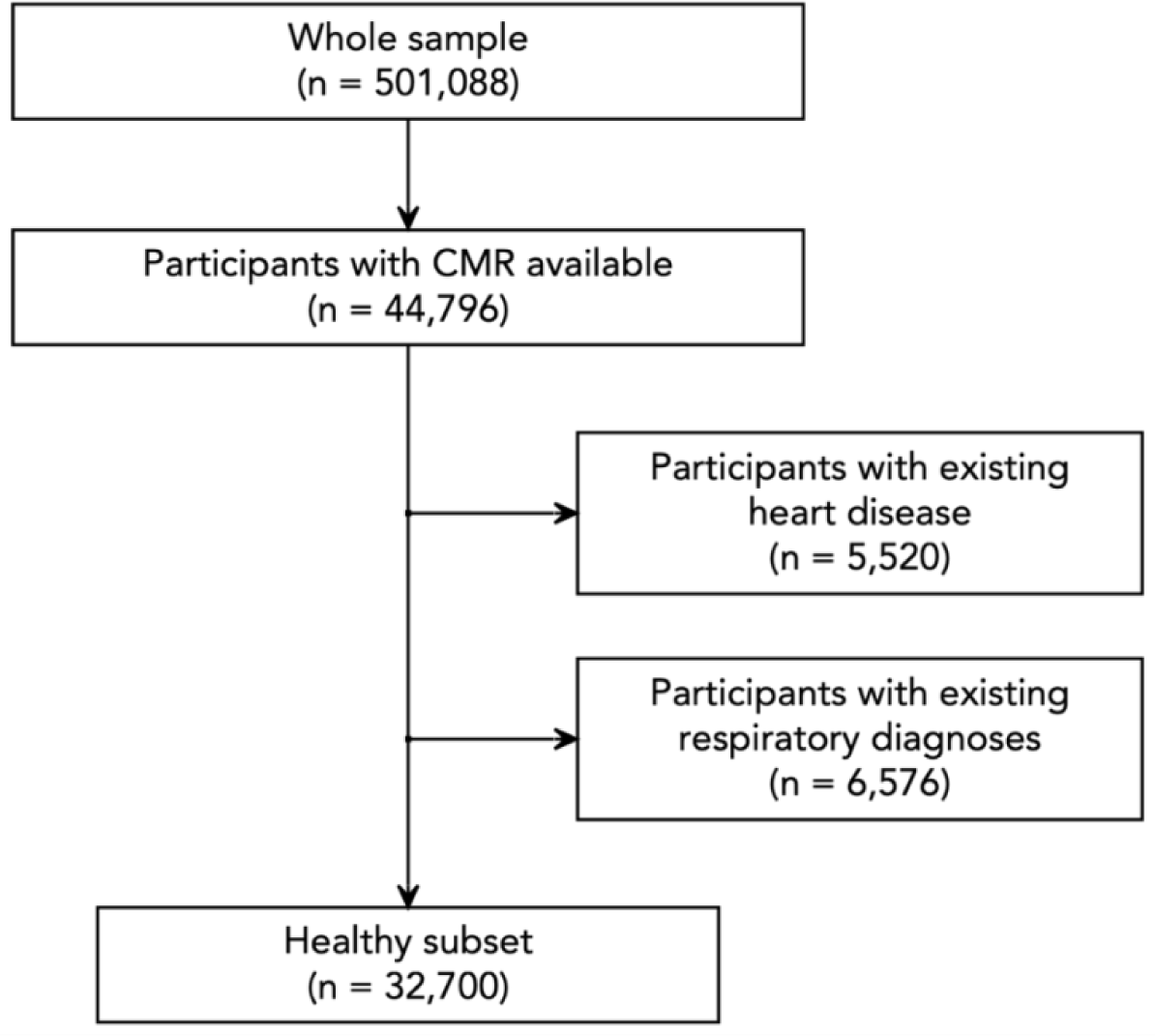
Study sample. CMR = cardiovascular magnetic resonance.

**Supplementary Table 1:**
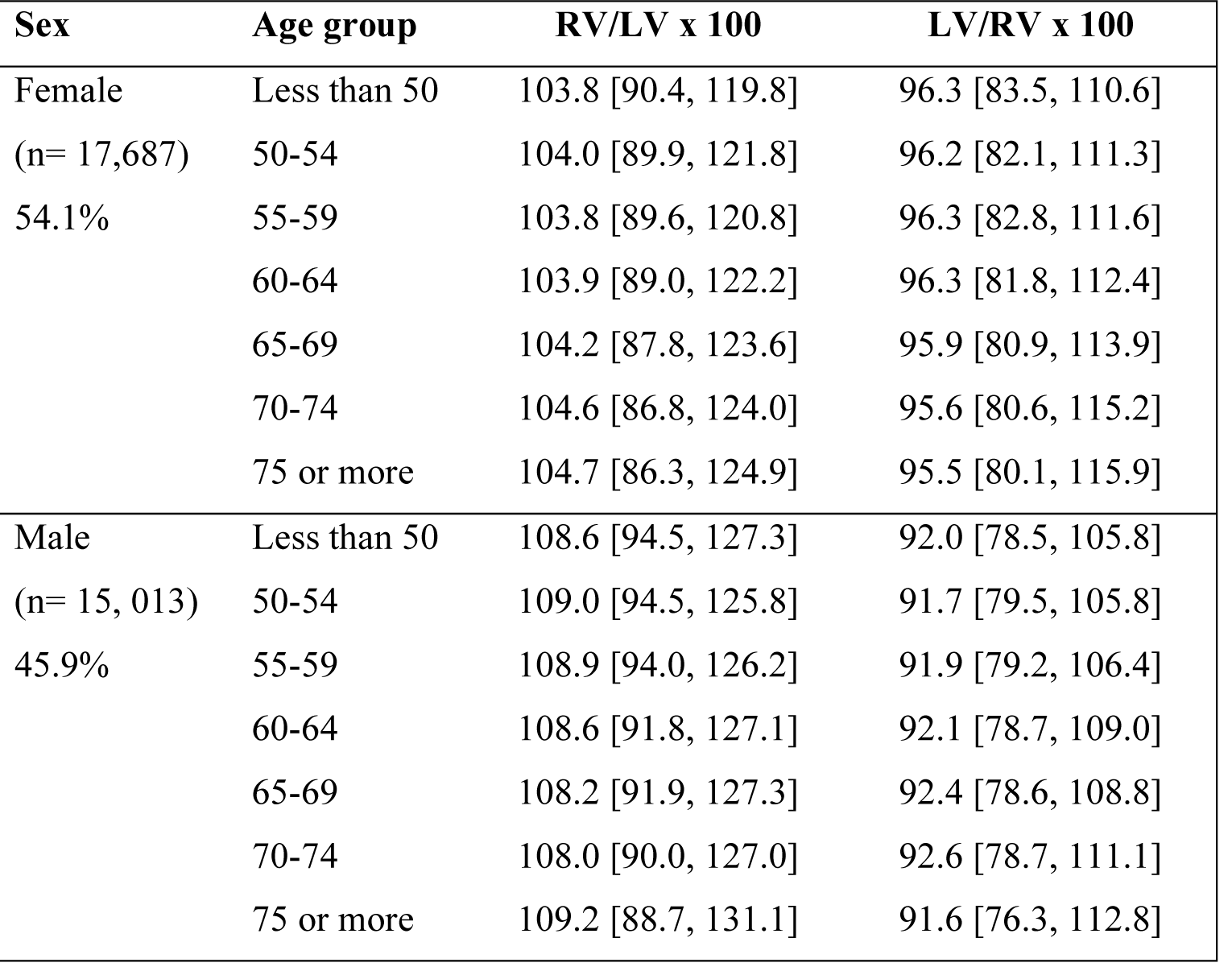
Ventricular ratio percentiles by sex and age. Entries are median ventricular asymmetry in the healthy subset, by sex and age [5^th^ percentile, 95^th^ percentile]. RV = right ventricular end-diastolic volume, LV = left ventricular end-diastolic volume. Ratios can be calculated with either raw or indexed ventricular volumes.

**Supplementary Table 2:**
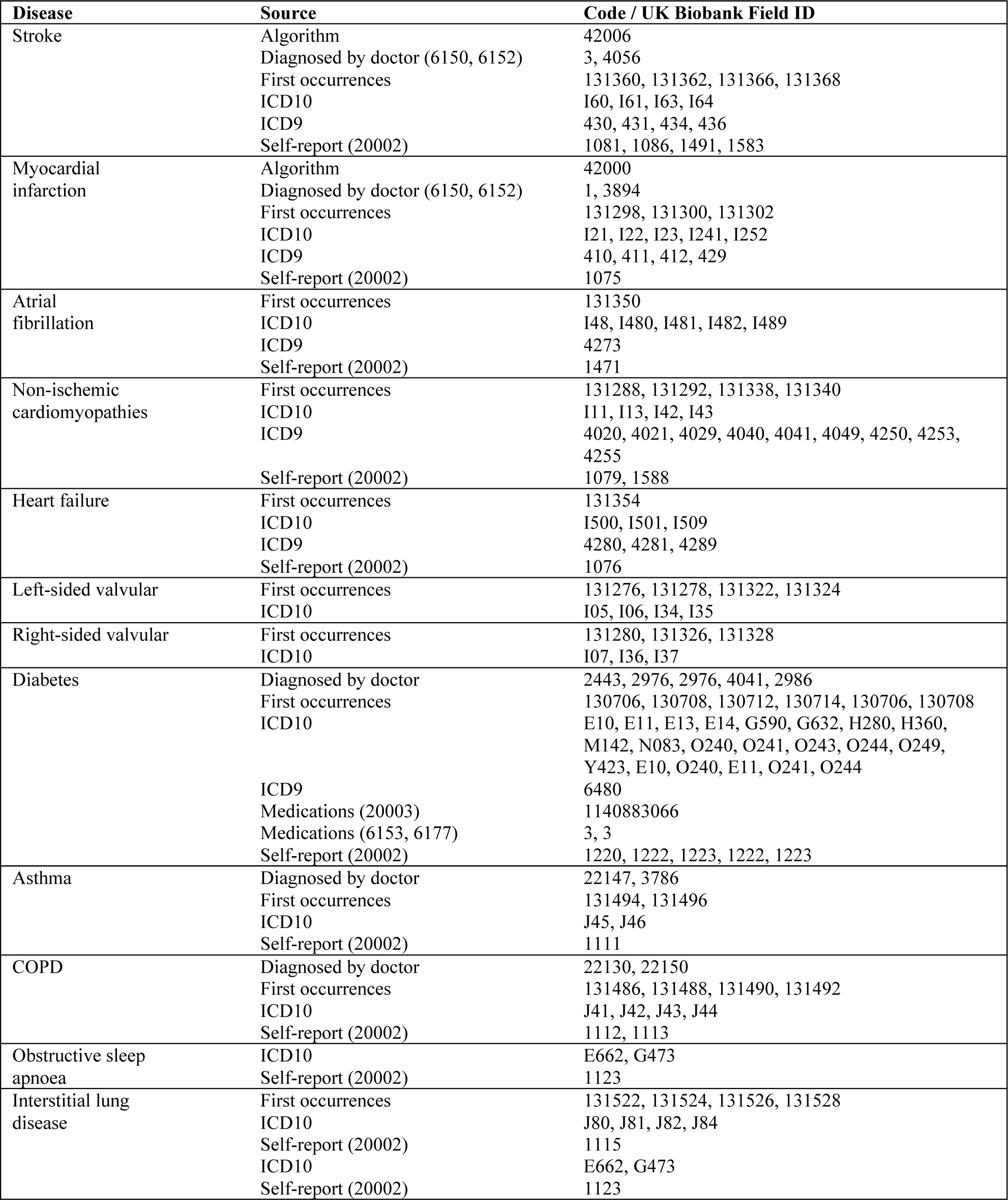
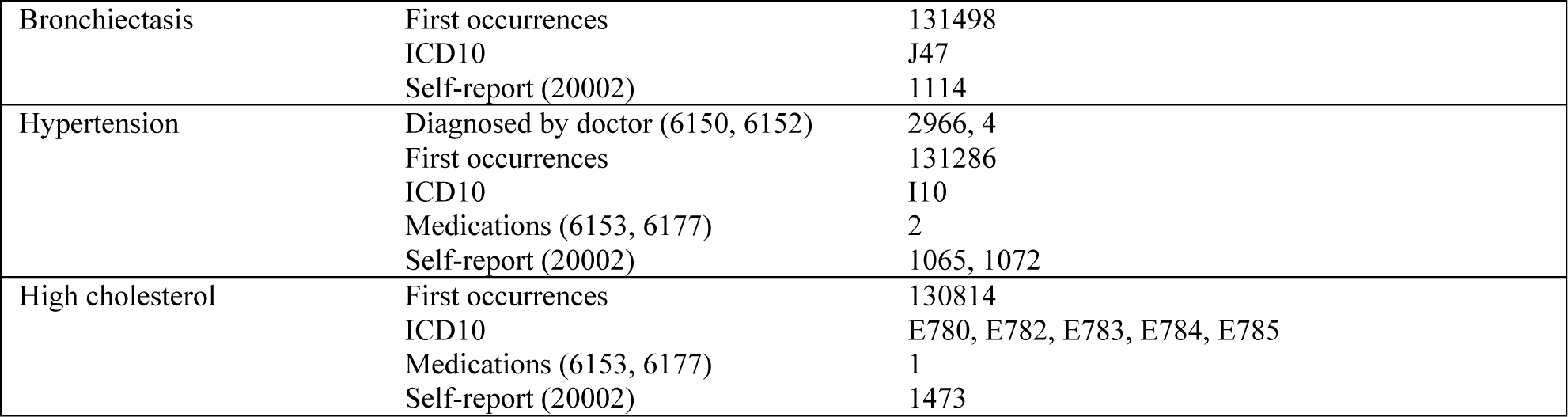
Disease definitions. ICD10 codes are drawn from fields 41270, 41280, 41234 and 41259; ICD9 codes are drawn from 41271 and 41281, Where a 3-digit code is given, this includes all 4-digit sub-codes, for example, E10 includes E100, E101 and E102.

**Supplementary Table 3:** Prevalent and Incident disease counts. COPD = chronic obstructive pulmonary disease, CVD = cardiovascular disease

| Condition | Whole sample<br>(n= 44,796) | Within<br>normal<br>symmetry<br>(n= 39,875) | RV dominant<br>(n= 2,263) | LV dominant<br>(n= 2,658) |
| --- | --- | --- | --- | --- |
| **Existing/prevalent at imaging** |  |  |  |  |
| Stroke | 914 (2.0%) | 778 (2.0%) | 49 (2.2%) | 87 (3.3%) |
| Asthma | 6,232 (13.9%) | 5,532 (13.9%) | 312 (13.8%) | 388 (14.6%) |
| COPD | 949 (2.1%) | 798 (2.0%) | 62 (2.7%) | 89 (3.3%) |
| Interstitial lung disease | 119 (0.3%) | 99 (0.2%) | 5 (0.2%) | 15 (0.6%) |
| Obstructive sleep apnoea | 569 (1.3%) | 507 (1.3%) | 21 (0.9%) | 41 (1.5%) |
| Bronchiectasis | 287 (0.6%) | 236 (0.6%) | 22 (1.0%) | 29 (1.1%) |
| Ischemic heart disease | 2,797 (6.2%) | 2,245 (5.6%) | 141 (6.2%) | 411 (15.5%) |
| Myocardial infarction | 1,148 (2.6%) | 855 (2.1%) | 40 (1.8%) | 253 (9.5%) |
| Atrial fibrillation | 1,358 (3.0%) | 1,129 (2.8%) | 97 (4.3%) | 132 (5.0%) |
| Non-ischemic cardiomyopathies | 108 (0.2%) | 72 (0.2%) | 2 (0.1%) | 34 (1.3%) |
| Right-sided valvular disorder<br>(tricuspid/pulmonary) | 61 (0.1%) | 46 (0.1%) | 11 (0.5%) | 4 (0.2%) |
| Left-sided valvular disorder<br>(mitral/aortic) | 401 (0.9%) | 290 (0.7%) | 15 (0.7%) | 96 (3.6%) |
| Multiple valve diseases | 124 (0.3%) | 91 (0.2%) | 6 (0.3%) | 27 (1.0%) |
| Heart failure | 285 (0.6%) | 186 (0.5%) | 17 (0.8%) | 82 (3.1%) |
| **Incident/ after imaging** |  |  |  |  |
| Stroke | 371 (0.8%) | 306 (0.8%) | 25 (1.1%) | 40 (1.5%) |
| Any CVD | 2,239 (5.0%) | 1,856 (4.7%) | 110 (4.9%) | 273 (10.3%) |
| Myocardial infarction | 502 (1.1%) | 427 (1.1%) | 21 (0.9%) | 54 (2.0%) |
| Atrial fibrillation | 958 (2.1%) | 759 (1.9%) | 53 (2.3%) | 146 (5.5%) |
| Non-ischemic cardiomyopathies | 102 (0.2%) | 66 (0.2%) | 3 (0.1%) | 33 (1.2%) |
| Right-sided valvular disorder<br>(tricuspid/pulmonary) | 81 (0.2%) | 69 (0.2%) | 7 (0.3%) | 5 (0.2%) |
| Left-sided valvular disorder<br>(mitral/aortic) | 343 (0.8%) | 238 (0.6%) | 12 (0.5%) | 93 (3.5%) |
| Heart failure | 421 (0.9%) | 266 (0.7%) | 23 (1.0%) | 132 (5.0%) |
| CVD mortality | 157 (0.4%) | 121 (0.3%) | 26 (1.0%) | 10 (0.4%) |
| All-cause mortality | 785 (1.8%) | 650 (1.6%) | 58 (2.6%) | 77 (2.9%) |
| Follow-up time (years) | 4.75 (± 1.5) | | | |

COPD = chronic obstructive pulmonary disease, CVD = cardiovascular disease \(Back to Results\)

**Supplementary Table 4:**
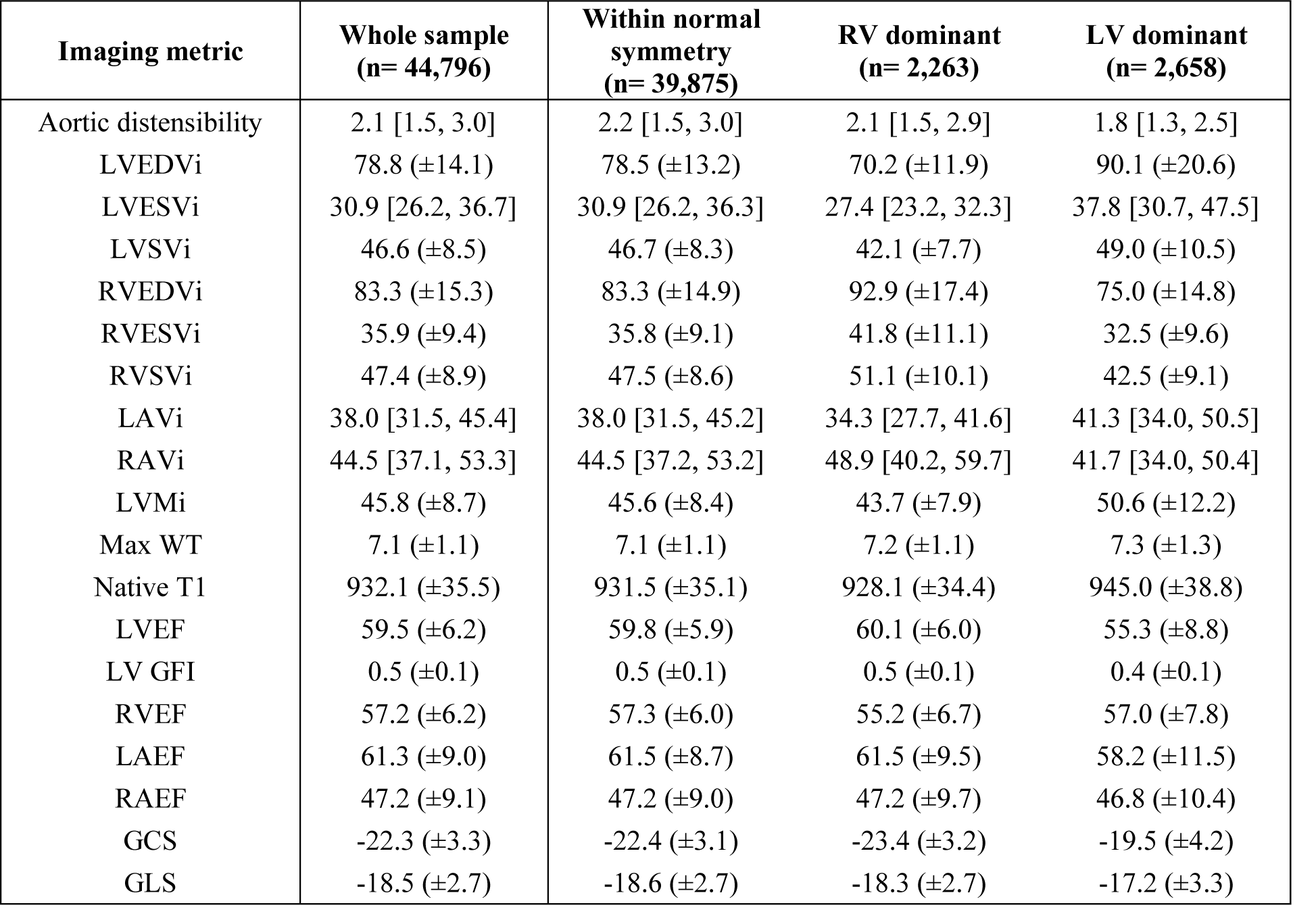
Imaging metric by asymmetry type. Entries are mean (± SD) or median [25^th^ percentile, 75^th^ percentile]. LVEDVi = left ventricular end-diastolic volume indexed to body surface area, LVESVi = left ventricular end-systolic volume indexed to body surface area, LVSVi = left ventricular stroke volume indexed to body surface area, RVEDVi = right ventricular end-diastolic volume indexed to body surface area, RVESVi = right ventricular end-systolic volume indexed to body surface area, RVSVi = right ventricular stroke volume indexed to body surface area, LAVi = maximum left atrial volume indexed to body surface area, RAVi = maximum right atrial volume indexed to body surface area, LVMi = left ventricular mass indexed to body surface area, WT= wall thickness, LVEF = left ventricular ejection fraction, LV GFI = left ventricular global function index, RVEF = right ventricular ejection fraction, LAEF= left atrial ejection fraction, RAEF= left atrial ejection fraction, GCS = global circumferential strain, GLS = global longitudinal strain.

| Imaging metric | Whole sample<br>(n= 44,796) | Within normal<br>symmetry<br>(n= 39,875) | RV dominant<br>(n= 2,263) | LV dominant<br>(n= 2,658) |
| --- | --- | --- | --- | --- |
| Aortic distensibility | 2.1 [1.5, 3.0] | 2.2 [1.5, 3.0] | 2.1 [1.5, 2.9] | 1.8 [1.3, 2.5] |
| LVEDVi | 78.8 (±14.1) | 78.5 (±13.2) | 70.2 (±11.9) | 90.1 (±20.6) |
| LVESVi | 30.9 [26.2, 36.7] | 30.9 [26.2, 36.3] | 27.4 [23.2, 32.3] | 37.8 [30.7, 47.5] |
| LVSVi | 46.6 (±8.5) | 46.7 (±8.3) | 42.1 (±7.7) | 49.0 (±10.5) |
| RVEDVi | 83.3 (±15.3) | 83.3 (±14.9) | 92.9 (±17.4) | 75.0 (±14.8) |
| RVESVi | 35.9 (±9.4) | 35.8 (±9.1) | 41.8 (±11.1) | 32.5 (±9.6) |
| RVSVi | 47.4 (±8.9) | 47.5 (±8.6) | 51.1 (±10.1) | 42.5 (±9.1) |
| LAVi | 38.0 [31.5, 45.4] | 38.0 [31.5, 45.2] | 34.3 [27.7, 41.6] | 41.3 [34.0, 50.5] |
| RAVi | 44.5 [37.1, 53.3] | 44.5 [37.2, 53.2] | 48.9 [40.2, 59.7] | 41.7 [34.0, 50.4] |
| LVMi | 45.8 (±8.7) | 45.6 (±8.4) | 43.7 (±7.9) | 50.6 (±12.2) |
| Max WT | 7.1 (±1.1) | 7.1 (±1.1) | 7.2 (±1.1) | 7.3 (±1.3) |
| Native T1 | 932.1 (±35.5) | 931.5 (±35.1) | 928.1 (±34.4) | 945.0 (±38.8) |
| LVEF | 59.5 (±6.2) | 59.8 (±5.9) | 60.1 (±6.0) | 55.3 (±8.8) |
| LV GFI | 0.5 (±0.1) | 0.5 (±0.1) | 0.5 (±0.1) | 0.4 (±0.1) |
| RVEF | 57.2 (±6.2) | 57.3 (±6.0) | 55.2 (±6.7) | 57.0 (±7.8) |
| LAEF | 61.3 (±9.0) | 61.5 (±8.7) | 61.5 (±9.5) | 58.2 (±11.5) |
| RAEF | 47.2 (±9.1) | 47.2 (±9.0) | 47.2 (±9.7) | 46.8 (±10.4) |
| GCS | -22.3 (±3.3) | -22.4 (±3.1) | -23.4 (±3.2) | -19.5 (±4.2) |
| GLS | -18.5 (±2.7) | -18.6 (±2.7) | -18.3 (±2.7) | -17.2 (±3.3) |

**Supplementary Table 5:** Ventricular asymmetry by clinical factors. Entries are odds ratios for ventricular asymmetry associated with each clinical feature, along with 95% confidence intervals, and p-values from unadjusted logistic regression models. An asterisk (*) indicates where p-values are significant after multiple testing adjustment with a 5% false discovery rate.

| Feature | RV dominant | LV dominant | Mildly LV dominant |
| --- | --- | --- | --- |
| Age 70 or more | 1.45*<br>[1.33, 1.59]<br>1.17x10<sup>-16</sup> | 1.83*<br>[1.69, 1.98]<br>6.28x10<sup>-49</sup> | 1.37*<br>[1.30, 1.45]<br>5.98x10<sup>-29</sup> |
| Male sex | 0.98<br>[0.90, 1.07]<br>0.6531 | 1.16*<br>[1.08, 1.26]<br>1.54x10<sup>-4</sup> | 1.00<br>[0.95, 1.05]<br>0.9310 |
| Ethnicities other than White | 1.07<br>[0.84, 1.37]<br>0.5660 | 0.78<br>[0.60, 1.01]<br>0.0551 | 0.80*<br>[0.68, 0.95]<br>0.0088 |
| Above median UK deprivation (more deprived) | 0.98<br>[0.89, 1.08]<br>0.7243 | 1.04<br>[0.95, 1.13]<br>0.4515 | 1.05<br>[0.99, 1.11]<br>0.1174 |
| Obesity (BMI > 30 kg/m2) | 0.85*<br>[0.75, 0.95]<br>0.0052 | 1.13*<br>[1.02, 1.25]<br>0.0153 | 1.10*<br>[1.02, 1.17]<br>0.0073 |
| Waist-hip ratio greater than 1 | 1.11<br>[0.94, 1.30]<br>0.2170 | 1.33*<br>[1.16, 1.53]<br>5.99x10<sup>-5</sup> | 1.26*<br>[1.14, 1.38]<br>3.88x10<sup>-6</sup> |
| Current smoker | 0.77<br>[0.59, 1.00]<br>0.0518 | 1.39*<br>[1.14, 1.70]<br>0.0013 | 1.40*<br>[1.22, 1.60]<br>9.26x10<sup>-7</sup> |
| Previous smoker | 0.95<br>[0.87, 1.04]<br>0.3043 | 1.28*<br>[1.18, 1.39]<br>2.05x10<sup>-9</sup> | 1.16*<br>[1.10, 1.23]<br>1.32x10<sup>-7</sup> |
| SBP above 140 mmHg | 0.83*<br>[0.77, 0.91]<br>4.10x10<sup>-5</sup> | 1.68*<br>[1.55, 1.82]<br>7.26x10<sup>-38</sup> | 1.46*<br>[1.39, 1.54]<br>2.03x10<sup>-46</sup> |
| Family history of heart disease | 1.00<br>[0.92, 1.09]<br>0.9610 | 1.07<br>[0.99, 1.16]<br>0.0735 | 1.07*<br>[1.01, 1.12]<br>0.0184 |
| Physically active (>600 METs/week) | 0.88*<br>[0.79, 0.98]<br>0.0238 | 0.83*<br>[0.75, 0.91]<br>2.28x10<sup>-4</sup> | 0.94<br>[0.87, 1.00]<br>0.0654 |

**Supplementary Table 6:**
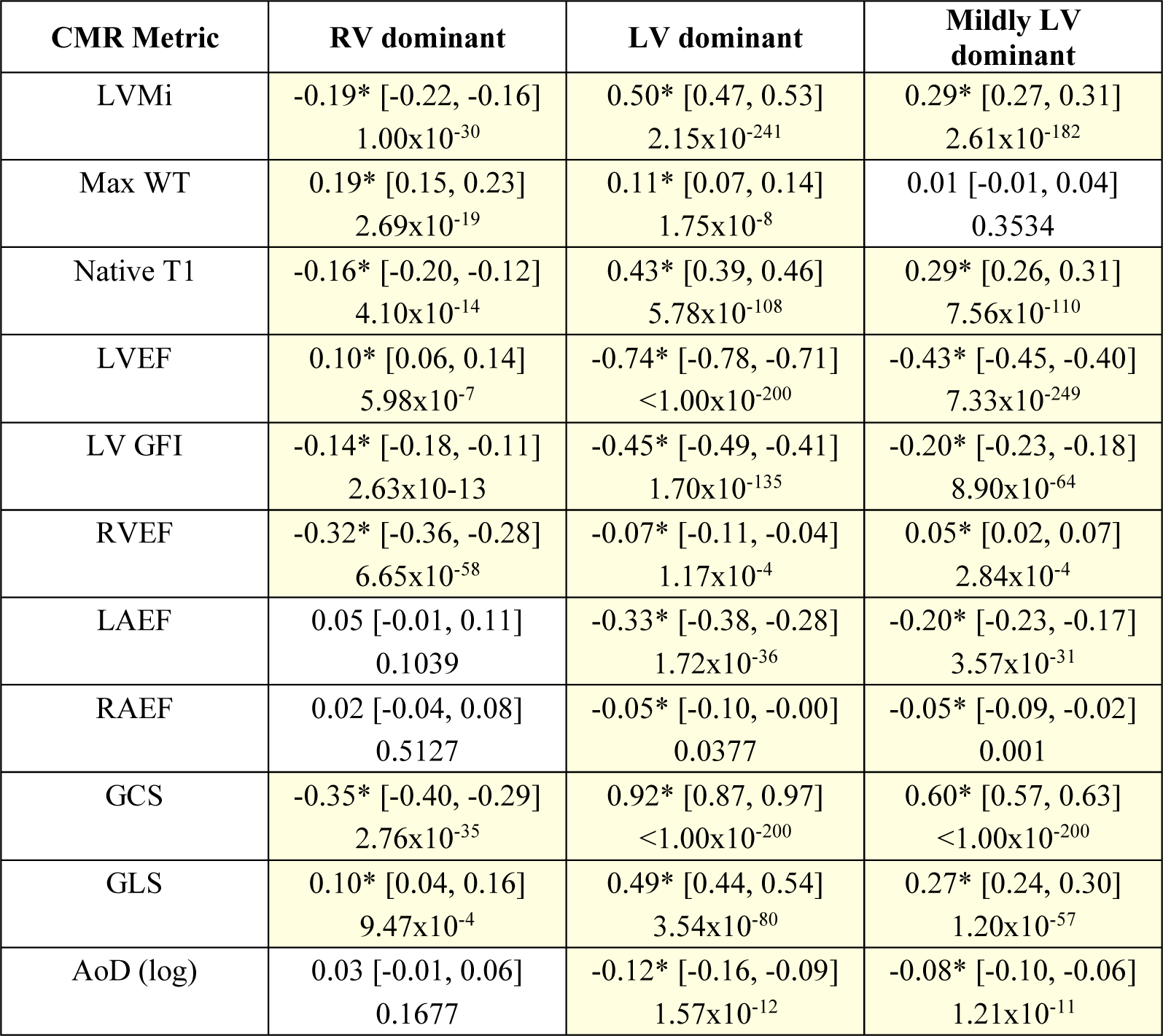
Associations between ventricular asymmetry and other CMR metrics. Entries are standardised beta coefficients with 95% confidence intervals and p-values for differences in CMR measures associated with each type of ventricular asymmetry compared to hearts within the normal symmetry range. Each cell is from a separate linear regression model, relating symmetry category to CMR metric, adjusted by age, sex, body mass index, systolic blood pressure, smoking, Townsend deprivation index, clinically diagnosed hypertension, diabetes and high cholesterol. An asterisk (*) indicates p-values that are significant after multiple testing adjustment with a false discovery rate of 5%.

| CMR Metric | RV dominant | LV dominant | Mildly LV dominant |
| --- | --- | --- | --- |
| LVMi | -0.19* [-0.22, -0.16]<br>1.00x10<sup>-30</sup> | 0.50* [0.47, 0.53]<br>2.15x10<sup>-241</sup> | 0.29* [0.27, 0.31]<br>2.61x10<sup>-182</sup> |
| Max WT | 0.19* [0.15, 0.23]<br>2.69x10<sup>-19</sup> | 0.11* [0.07, 0.14]<br>1.75x10<sup>-8</sup> | 0.01 [-0.01, 0.04]<br>0.3534 |
| Native T1 | -0.16* [-0.20, -0.12]<br>4.10x10<sup>-14</sup> | 0.43* [0.39, 0.46]<br>5.78x10<sup>-108</sup> | 0.29* [0.26, 0.31]<br>7.56x10<sup>-110</sup> |
| LVEF | 0.10* [0.06, 0.14]<br>5.98x10<sup>-7</sup> | -0.74* [-0.78, -0.71]<br><1.00x10<sup>-200</sup> | -0.43* [-0.45, -0.40]<br>7.33x10<sup>-249</sup> |
| LV GFI | -0.14* [-0.18, -0.11]<br>2.63x10<sup>-13</sup> | -0.45* [-0.49, -0.41]<br>1.70x10<sup>-135</sup> | -0.20* [-0.23, -0.18]<br>8.90x10<sup>-64</sup> |
| RVEF | -0.32* [-0.36, -0.28]<br>6.65x10<sup>-58</sup> | -0.07* [-0.11, -0.04]<br>1.17x10<sup>-4</sup> | 0.05* [0.02, 0.07]<br>2.84x10<sup>-4</sup> |
| LAEF | 0.05 [-0.01, 0.11]<br>0.1039 | -0.33* [-0.38, -0.28]<br>1.72x10<sup>-36</sup> | -0.20* [-0.23, -0.17]<br>3.57x10<sup>-31</sup> |
| RAEF | 0.02 [-0.04, 0.08]<br>0.5127 | -0.05* [-0.10, -0.00]<br>0.0377 | -0.05* [-0.09, -0.02]<br>0.001 |
| GCS | -0.35* [-0.40, -0.29]<br>2.76x10<sup>-35</sup> | 0.92* [0.87, 0.97]<br><1.00x10<sup>-200</sup> | 0.60* [0.57, 0.63]<br><1.00x10<sup>-200</sup> |
| GLS | 0.10* [0.04, 0.16]<br>9.47x10<sup>-4</sup> | 0.49* [0.44, 0.54]<br>3.54x10<sup>-80</sup> | 0.27* [0.24, 0.30]<br>1.20x10<sup>-57</sup> |
| AoD (log) | 0.03 [-0.01, 0.06]<br>0.1677 | -0.12* [-0.16, -0.09]<br>1.57x10<sup>-12</sup> | -0.08* [-0.10, -0.06]<br>1.21x10<sup>-11</sup> |

**Supplementary Table 7:**
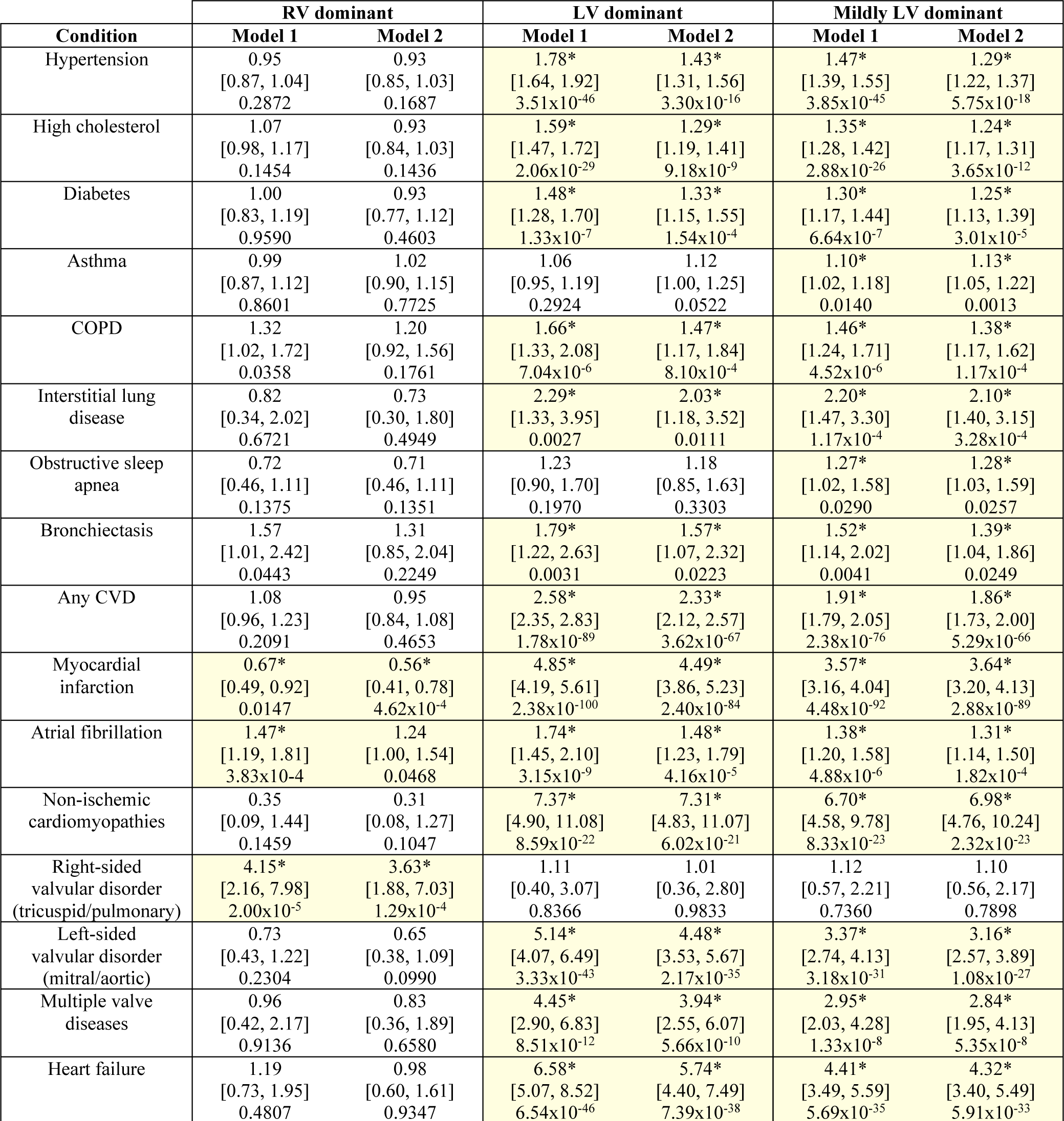
Associations between ventricular asymmetry and existing disease. Entries are odds ratio for ventricular asymmetry associated with the presence of each existing diagnosis, along with 95% confidence intervals and p-values, calculated with logistic regression. Model 1 is the crude, unadjusted association. Model 2 is adjusted by age, sex, body mass index, systolic blood pressure and Townsend deprivation index. An asterisk (*) indicates p-values that are significant after multiple testing adjustment with a false discovery rate of 5%.

**Supplementary Table 8:** Associations between ventricular ratio and incident disease. Entries are hazard ratios for each disease associated with each volume asymmetry type, with 95% confidence intervals and p-values from Cox proportional hazard regression models. Model 1= crude association, Model 2 includes adjustment by age, sex, systolic blood pressure, body mass index, smoking, Townsend deprivation index, hypertension, high cholesterol, diabetes. Model 3 includes Model 2 covariates plus left ventricular mass index, right ventricular ejection fraction and left ventricular ejection fraction. Two-tailed significance was adjusted for multiple testing with a false discovery rate of 5%.

|  | RV dominates |  |  | LV dominates |  |  | Mildly LV dominant |  |  |
| --- | --- | --- | --- | --- | --- | --- | --- | --- | --- |
|  | Model 1 | Model 2 | Model 3 | Model 1 | Model 2 | Model 3 | Model 1 | Model 2 | Model 3 |
| Stroke | 1.44<br>[0.96, 2.16]<br>0.0773 | 1.36<br>[0.91, 2.05]<br>0.1378 | 1.44<br>[0.95, 2.17]<br>0.0850 | 1.95*<br>[1.40, 2.70]<br>6.97x10<sup>-5</sup> | 1.50*<br>[1.08, 2.09]<br>0.0159 | 1.12<br>[0.77, 1.62]<br>0.5492 | 1.46*<br>[1.14, 1.88]<br>0.0031 | 1.26<br>[0.98, 1.63]<br>0.0731 | 1.05<br>[0.80, 1.39]<br>0.7084 |
| Any CVD | 1.04<br>[0.85, 1.25]<br>0.7219 | 0.96<br>[0.79, 1.16]<br>0.6675 | 1.04<br>[0.85, 1.26]<br>0.7153 | 2.70*<br>[2.38, 3.06]<br>2.91x10<sup>-53</sup> | 2.23*<br>[1.96, 2.53]<br>3.86x10<sup>-34</sup> | 1.79*<br>[1.56, 2.06]<br>1.59x10<sup>-16</sup> | 1.77*<br>[1.60, 1.95]<br>6.72x10<sup>-29</sup> | 1.60*<br>[1.44, 1.77]<br>1.38x10<sup>-19</sup> | 1.38*<br>[1.24, 1.54]<br>2.75x10<sup>-9</sup> |
| Myocardial infarction | 0.86<br>[0.56, 1.33]<br>0.5037 | 0.80<br>[0.51, 1.26]<br>0.3373 | 0.89<br>[0.57, 1.40]<br>0.6094 | 2.08*<br>[1.57, 2.76]<br>3.81x10<sup>-7</sup> | 1.61*<br>[1.21, 2.14]<br>0.0011 | 1.28<br>[0.94, 1.75]<br>0.1163 | 1.77*<br>[1.44, 2.18]<br>6.44x10<sup>-8</sup> | 1.51*<br>[1.22, 1.86]<br>1.46x10<sup>-4</sup> | 1.31*<br>[1.05, 1.65]<br>0.0181 |
| Atrial fibrillation | 1.18<br>[0.89, 1.55]<br>0.2499 | 1.09<br>[0.83, 1.44]<br>0.5440 | 1.17<br>[0.88, 1.55]<br>0.2748 | 2.97*<br>[2.49, 3.54]<br>9.63x10<sup>-34</sup> | 2.24*<br>[1.87, 2.68]<br>1.24x10<sup>-18</sup> | 1.67*<br>[1.36, 2.05]<br>7.01x10<sup>-7</sup> | 2.04*<br>[1.76, 2.35]<br>3.67x10<sup>-22</sup> | 1.73*<br>[1.49, 2.00]<br>2.66x10<sup>-13</sup> | 1.43*<br>[1.22, 1.68]<br>9.69x10<sup>-6</sup> |
| Non-ischemic<br>cardiomyopathies | 0.61<br>[0.19, 1.94]<br>0.4058 | 0.58<br>[0.18, 1.83]<br>0.3508 | 0.93<br>[0.29, 2.98]<br>0.9089 | 7.73*<br>[5.10, 11.70]<br>4.39x10<sup>-22</sup> | 6.34*<br>[4.13, 9.74]<br>2.95x10<sup>-17</sup> | 2.43*<br>[1.41, 4.19]<br>0.0014 | 5.02*<br>[3.40, 7.40]<br>4.31x10<sup>-16</sup> | 4.53*<br>[3.05, 6.74]<br>7.98x10<sup>-14</sup> | 2.36*<br>[1.49, 3.74]<br>2.62x10<sup>-4</sup> |
| Right-sided valvular disorder<br>(tricuspid/pulmonary) | 1.89<br>[0.87, 4.11]<br>0.1071 | 1.75<br>[0.81, 3.82]<br>0.1571 | 1.82<br>[0.83, 4.02]<br>0.1363 | 1.05<br>[0.42, 2.60]<br>0.9157 | 0.65<br>[0.24, 1.78]<br>0.3994 | 0.49<br>[0.17, 1.43]<br>0.1902 | 0.80<br>[0.41, 1.54]<br>0.4980 | 0.61<br>[0.31, 1.23]<br>0.1716 | 0.51<br>[0.24, 1.06]<br>0.0724 |
| Left-sided valvular disorder<br>(mitral/aortic) | 0.71<br>[0.40, 1.26]<br>0.2423 | 0.60<br>[0.33, 1.09]<br>0.0957 | 0.71<br>[0.39, 1.30]<br>0.2657 | 6.22*<br>[4.90, 7.89]<br>3.66x10<sup>-51</sup> | 4.58*<br>[3.60, 5.84]<br>1.08x10<sup>-34</sup> | 3.43*<br>[2.59, 4.52]<br>4.15x10<sup>-18</sup> | 3.72*<br>[3.00, 4.62]<br>1.61x10<sup>-32</sup> | 3.09*<br>[2.48, 3.86]<br>1.06x10<sup>-23</sup> | 2.47*<br>[1.94, 3.14]<br>2.08x10<sup>-13</sup> |
| Heart failure | 1.16<br>[0.76, 1.76]<br>0.4968 | 1.04<br>[0.68, 1.58]<br>0.8713 | 1.45<br>[0.95, 2.22]<br>0.0883 | 7.63*<br>[6.21, 9.37]<br>2.37x10<sup>-83</sup> | 5.53*<br>[4.48, 6.83]<br>7.46x10<sup>-57</sup> | 2.57*<br>[1.98, 3.34]<br>1.46x10<sup>-12</sup> | 4.43*<br>[3.65, 5.37]<br>9.69x10<sup>-52</sup> | 3.61*<br>[2.96, 4.39]<br>1.70x10<sup>-37</sup> | 2.04*<br>[1.62, 2.55]<br>8.69x10<sup>-10</sup> |
| CVD mortality | 1.38<br>[0.72, 2.61]<br>0.3291 | 1.13<br>[0.58, 2.22]<br>0.7246 | 1.32<br>[0.67, 2.62]<br>0.4251 | 3.14*<br>[2.06, 4.79]<br>9.75x10<sup>-8</sup> | 2.27*<br>[1.48, 3.48]<br>1.69x10<sup>-4</sup> | 1.24<br>[0.74, 2.07]<br>0.4076 | 2.54*<br>[1.81, 3.55]<br>6.64x10<sup>-8</sup> | 2.08*<br>[1.48, 2.94]<br>3.05x10<sup>-5</sup> | 1.46<br>[0.99, 2.16]<br>0.0569 |
| All-cause mortality | 1.62*<br>[1.24, 2.11]<br>4.34x10<sup>-4</sup> | 1.35<br>[1.02, 1.78]<br>0.0353 | 1.41*<br>[1.07, 1.87]<br>0.0162 | 1.72*<br>[1.36, 2.18]<br>6.06x10<sup>-6</sup> | 1.35*<br>[1.06, 1.71]<br>0.0140 | 1.14<br>[0.88, 1.47]<br>0.3289 | 1.43*<br>[1.20, 1.70]<br>5.56x10<sup>-5</sup> | 1.25*<br>[1.04, 1.49]<br>0.0144 | 1.12<br>[0.93, 1.36]<br>0.2268 |

## Abbreviations list

AF: atrial fibrillation
AoD: aortic distensibility
CMR: cardiac magnetic resonance
COPD: chronic obstructive pulmonary disease
CVD: cardiovascular disease
GCS: global circumferential strain
GLS: global longitudinal strain.
IHD: ischemic heart disease
LAEF: left atrial ejection fraction
LAVi: left atrial volume index
LVEDVi: left ventricular end-diastolic volume indexed to body surface area
LVEF: left ventricular volume
LVESVi: left ventricular end-systolic volume index
LVGFI: left ventricular performance index
LVMi: left ventricular mass index
LVSVi: left ventricular stroke volume index
Max WT: maximum wall thickness
MI: myocardial infarction
RAEF: right atrial ejection fraction
RVEDVi: right ventricular end-diastolic volume indexed to body surface area
RVEF: right ventricular volume
RVESVi: right ventricular end-systolic volume index
RVSVi: right ventricular stroke volume index

